# Healthy ageing men have normal reproductive function but display germline-specific molecular changes

**DOI:** 10.1101/19006221

**Authors:** S. Laurentino, J.-F. Cremers, B. Horsthemke, F. Tüttelmann, K. Czeloth, M. Zitzmann, E. Pohl, S. Rahmann, C. Schröder, S. Berres, K. Redmann, C. Krallmann, S. Schlatt, S. Kliesch, J. Gromoll

**Affiliations:** Centre of Reproductive Medicine and Andrology, University of Münster, Albert-Schweitzer Campus 1, Building D11, 48149 Münster, Germany; Institute of Human Genetics, University of Duisburg-Essen, University Hospital Essen, Hufelandstr. 55, 45147 Essen, Germany; Institute of Human Genetics, University of Münster, Vesaliusweg 12-14, 48149 Münster, Germany; Genome Informatics, University of Duisburg-Essen, University Hospital Essen, Hufelandstr. 55, 45147 Essen, Germany

**Author notes:** Shared last-authorship (S. Kliesch, J. Gromoll).

## Abstract

Children of older fathers have higher risk for certain diseases. Nevertheless, how ageing specifically affects male germ cells is so far not completely understood. In a cohort of 197 healthy men (18-84 years), we found that semen and reproductive parameters remained normal over six decades. Along with an age-dependent increase in telomere length in sperm (r=0.41, p>0.001), we found accelerated DNA fragmentation, more prominent after the sixth decate of life, and with around 60% of men older than 66 showing abnormal levels of DNA breaks. At the epigenetic level, by whole genome bisulfite sequencing we identified 236 sperm-specific differentially methylated regions between the youngest and oldest group, affecting mostly regions associated with homeobox genes and nervous system development. Therefore, we propose that during ageing, male germ cells are affected by an intrinsic and specific ageing process, distinguishable from the soma. These age-dependent changes might have consequences for fertility and offspring of older men.

## Introduction

In developed countries, couples postpone parenthood mainly for socio-political and cultural reasons (1–3). The reproductive implications of delaying fatherhood, such as a decline of sperm parameters (e.g. sperm count, motility, and morphology) and disruptions in reproductive hormone secretion (4), have long been undervalued. In fact, if and how ageing itself causes a decline in men’s reproductive health is unclear because selection criteria for clinical studies are inconsistent and some studies indicate only a subtle change during ageing (5, 6) which might in part be due to unhealthy lifestyles or age-related somatic diseases (7). Observational studies have indicated that the partners of elderly men take longer to become pregnant and that these pregnancies have higher rates of spontaneous abortion (8–10). Recently, advanced paternal age has been shown to have adverse consequences for maternal and offspring health (11). In spite of this, knowledge of how ageing specifically affects the quality of male germ cells remains limited.

The increasing number of pre-meiotic cell divisions in the human male germline leads to an increase in *de novo* mutation rates in children of older men (12, 13). In line with this, there is an association between paternal age and incidence of so called paternal age effect (PAE) disorders (14). Still, the mechanism by which PAE arise is not completely understood. Among the hypotheses put forward to explain the detrimental effects of age on male fertility and offspring health, epigenetic changes to the DNA stand out. Changes in DNA methylation accumulate in the male germline with age (15–17), however only a small portion of all CpG sites in the genome have so far been interrogated.

Our unique cross-sectional observation study on healthy ageing men [Fertility and Ageing in Healthy Men (FAMe)] was conducted in order to exclude the effects of age-related mobidities and evaluate the impacts of age to male reproductive health. Our ultimate goal was to identify the fundamental processes involved in male germline ageing which are caused exclusively by age.

## Results

### Reproductive parameters in healthy ageing men

We evaluated reproductive parameters in 197 healthy men, who were stratified into six age groups (Table 1). We did not observe any age-related changes in testicular volume or semen parameters beyond or below the normal ranges given by the WHO (18). Subtle, but significant age-dependent processes such as a decrease of ejaculate volume, changes in sperm motility, and a decrease in markers for accessory gland function, e.g. zinc were clearly visible, however all changes remained within the given physiological range (Table 2).

**Table 1.** Age distribution of the FAME cohort

| Group | Age Range (years) | N |
| --- | --- | --- |
| 1 | 18-25 | 34 |
| 2 | 26-35 | 36 |
| 3 | 36-45 | 28 |
| 4 | 46-55 | 39 |
| 5 | 56-65 | 36 |
| 6 | ≥66 | 24 |

**Table 2.** Semen parameters across age groups

|  | Group 1<br>n=34<br>18-25<br>years | Group 2<br>n=36<br>26-35<br>years | Group 3<br>n=28<br>36-45<br>years | Group 4<br>n=39<br>46-55<br>years | Group 5<br>n=36<br>56-65<br>years | Group 6<br>n= 24<br>>66<br>years | Correlation<br>with age |
| --- | --- | --- | --- | --- | --- | --- | --- |
| Testicular Vol.<br>(>15cm<sup>3</sup>) | 43.2<br>(13.4) | 41.5<br>(13.8) | 42.3<br>(15.7) | 39.1<br>(12.5) | 42.7<br>(12.9) | 39.6<br>(13.4) | -0.05 (0.51) |
| Ejaculate Vol.<br>(≥1.5ml) | 4.2 (1.7) | 4.6 (2.3) | 3.7<br>(1.8) | 3.2 (1.5) | 2.9 (1.5) | 2.0 (1.2) | -0.41<br>(3.86x10<sup>-9</sup>) |
| Sperm Conc.<br>(≥15 mill/ml) | 39.5<br>(37.3) | 47.6<br>(37.8) | 34.5<br>(30.7) | 53.5<br>(45.1) | 52.7<br>(50.1) | 62.9<br>(43.8) | 0.08 (0.29) |
| Total Sperm<br>Counts<br>(≥39 Mill) | 142.3<br>(116.2) | 206.5<br>(179.5) | 118.7<br>(86.0) | 165.4<br>(160.3) | 139.1<br>(140.5) | 122.3<br>(120.8) | -0.14 (0.06) |
| Prog Motility<br>(≥32%) | 55.2<br>(15.1) | 57.9<br>(12.5) | 47.8<br>(16.4) | 49.8<br>(16.2) | 43.4<br>(18.7) | 35.6<br>(21.1) | -0.39<br>(1.68x10<sup>-8</sup>) |
| Norm.<br>Morphology<br>(≥4%) | 4.9 (1.7) | 4.7 (1.8) | 4.6<br>(2.0) | 5.2 (2.1) | 4.4 (2.2) | 5.1 (2.1) | 0.01 (0.85) |
| Glucosidase<br>(>20 mU/Ejac.) | 87.0<br>(56.4) | 131.4<br>(76.3) | 80.3<br>(46.8) | 101.3<br>(76.0) | 120.4<br>(90.9) | 113.3<br>(70.9) | 0.09 (0.24) |
| Fructose (>13<br>μmol/Ejac.) | 64.2<br>(28.6) | 70.5<br>(54.1) | 48.3<br>(30.3) | 39.1<br>(25.9) | 35.0<br>(30.3) | 19.2<br>(19.5) | -0.47<br>(1.18x10<sup>-11</sup>) |
| Zinc (>2.4<br>μmol/Ejac) | 6.8 (4.3) | 8.8 (6.4) | 7.5<br>(5.1) | 6.7 (3.8) | 5.8 (4.6) | 6.8 (6.0) | -0.13 (0.09) |
| pH (≥7.2) | 8.1 (0.3) | 8.1 (0.3) | 8.3<br>(0.3) | 8.2 (0.3) | 8.4 (0.5) | 8.2 (0.3) | 0.19 (0.009) |
| Non progressive<br>motile (%) | 12.1 (6.8) | 10.3<br>(3.2) | 14.5<br>(8.7) | 9.7 (4.4) | 9.7 (5.9) | 11.1<br>(7.5) | -0.07 (0.34) |
| Immotile (%) | 32.7<br>(10.547) | 31.9<br>(9.0) | 37.9<br>(15.9) | 40.5<br>(17.2) | 41.8<br>(17.4) | 51.6<br>(21.2) | 0.37<br>(1.05x10<sup>-7</sup>) |
| Head defects (%) | 95.0 (3.5) | 95.4<br>(2.7) | 96.0<br>(2.0) | 95.3<br>(2.1) | 96.1<br>(2.1) | 95.7<br>(2.0) | 0.05 (0.53) |
| Midpiece defects<br>(%) | 54.4 (7.6) | 53.8<br>(7.4) | 54.6<br>(7.8) | 53.0<br>(6.7) | 53.0<br>(8.9) | 54.6<br>(7.0) | -0.01 (0.89) |
| Tail defects (%) | 15.6 (4.5) | 15.7<br>(4.9) | 15.4<br>(4.1) | 15.6<br>(5.1) | 16.2<br>(4.8) | 15.2<br>(4.3) | -0.02 (0.79) |
| Agglutination | 3.2 (7.2) | 5.8<br>(13.1) | 3.2<br>(10.1) | 3.6 (6.6) | 5.6<br>(13.3) | 9.1<br>(19.7) | 0.03 (0.69) |
| MAR IgG (<50%) | 5.9 (14.0) | 3.2 (8.3) | 8.0<br>(22.0) | 10.7<br>(23.0) | 8.2<br>(12.7) | 12.1<br>(25.3) | 0.15 (0.047) |
| MAR IgA (<50%) | 1.9 (6.2) | 1.1 (3.6) | 3.1<br>(8.5) | 5.7<br>(12.3) | 4.5 (7.8) | 6.7<br>(20.5) | 0.16 (0.04) |
| Vitality (>58%) | 71.2<br>(11.5) | 70.5<br>(11.4) | 66.0<br>(14.2) | 69.3<br>(11.4) | 60.6<br>(15.8) | 59.3<br>(15.9) | -0.31<br>(1.99x10<sup>-5</sup>) |
| Round cells<br>(mill/ml) | 1.3 (1.6) | 1.4 (1.8) | 1.2<br>(1.2) | 1.2 (1.3) | 1.6 (2.0) | 2.3 (3.9) | 0.05 (0.54) |
| Leucocytes (<1<br>mill/ml) | 0.4 (1.5) | 0.2 (0.7) | 0.2<br>(0.4) | 0.2 (0.4) | 0.5 (1.4) | 0.6 (1.2) | 0.05 (0.47) |

Normal ranges are indicated for each parameter. Results are shown as mean (SD) for each age group and Spearman's rank correlations with age as $\rho$ (p-value).

Similar observations were made regarding reproductive endocrine parameters. Although there was a slight age-dependent increase in FSH, LH, and SHBG and a decrease in free testosterone, all hormone levels stayed within the normal range (Table 3). A similar scenario was observed when evaluating anthropometric parameters and questionnaires for (sexual) wellbeing (Table 4).

**Table 3.** Hormonal profiles of the men in the study cohort

|  | Group 1<br>n=34<br>18-25<br>years | Group 2<br>n=36<br>26-35<br>years | Group 3<br>n=28<br>36-45<br>years | Group 4<br>n=39<br>46-55<br>years | Group 5<br>n=36<br>56-65<br>years | Group 6<br>n= 24<br>>66 years | Correlation<br>with age |
| --- | --- | --- | --- | --- | --- | --- | --- |
| FSH (2-10 U/l) | 3.0 (1.9) | 3.4 (1.5) | 4.0 (2.0) | 4.1 (2.4) | 5.2 (5.9) | 6.1 (3.0) | 0.36<br>($2.08 \times 10^{-7}$) |
| LH (1-7 U/l) | 3.0 (0.9) | 3.2 (1.1) | 3.5 (1.5) | 3.0 (1.1) | 2.8 (1.2) | 4.0 (1.7) | 0.02 (0.81) |
| Total T (>12 nmol/l) | 23.9 (7.3) | 23.8 (6.7) | 21.2 (6.8) | 19.4 (6.2) | 20.8 (6.2) | 24.1 (9.3) | -0.12 (0.09) |
| SHBG (11-71 nmol/l) | 38.2 (12.8) | 36.6 (12.8) | 37.2 (11.9) | 39.7 (13.1) | 47.3 (17.7) | 58.8 (19.3) | 0.32<br>($3.56 \times 10^{-6}$) |
| Free T (>250 pmol/l) | 494.4 (128.5) | 501.5 (103.8) | 435.8 (138.7) | 369.0 (111.3) | 361.2 (84.9) | 361.3 (117.4) | -0.47<br>($6.89 \times 10^{-12}$) |
| DHT (0.5-2.0 nmol/l) | 0.9 (0.2) | 0.8 (0.2) | 0.8 (0.2) | 0.8 (0.3) | 0.8 (0.3) | 1.0 (0.4) | -0.004 (0.95) |
| Estradiol (<250 pmol/l) | 89.0 (31.3) | 86.8 (26.9) | 85.6 (34.8) | 78.1 (28.8) | 88.5 (32.2) | 98.1 (33.4746) | 0.01 (0.87) |
| Prolactin (<500 mU/l) | 297.0 (165.0) | 303.0 (204.4) | 225.5 (97.2) | 189.4 (118.2) | 165.3 (92.6) | 199.4 (56.9) | -0.38<br>($3.43 \times 10^{-8}$) |
| PSA (<4 ug/l) | 0.5 (0.2) | 0.6 (0.3) | 0.7 (0.5) | 0.9 (0.8) | 1.5 (1.3) | 1.9 (1.7) | 0.45<br>($2.70 \times 10^{-11}$) |

Normal ranges are indicated for each parameter. Results are shown as mean (SD) for each age group and Spearman's rank correlations with age as p (p-value).

**Table 4.**
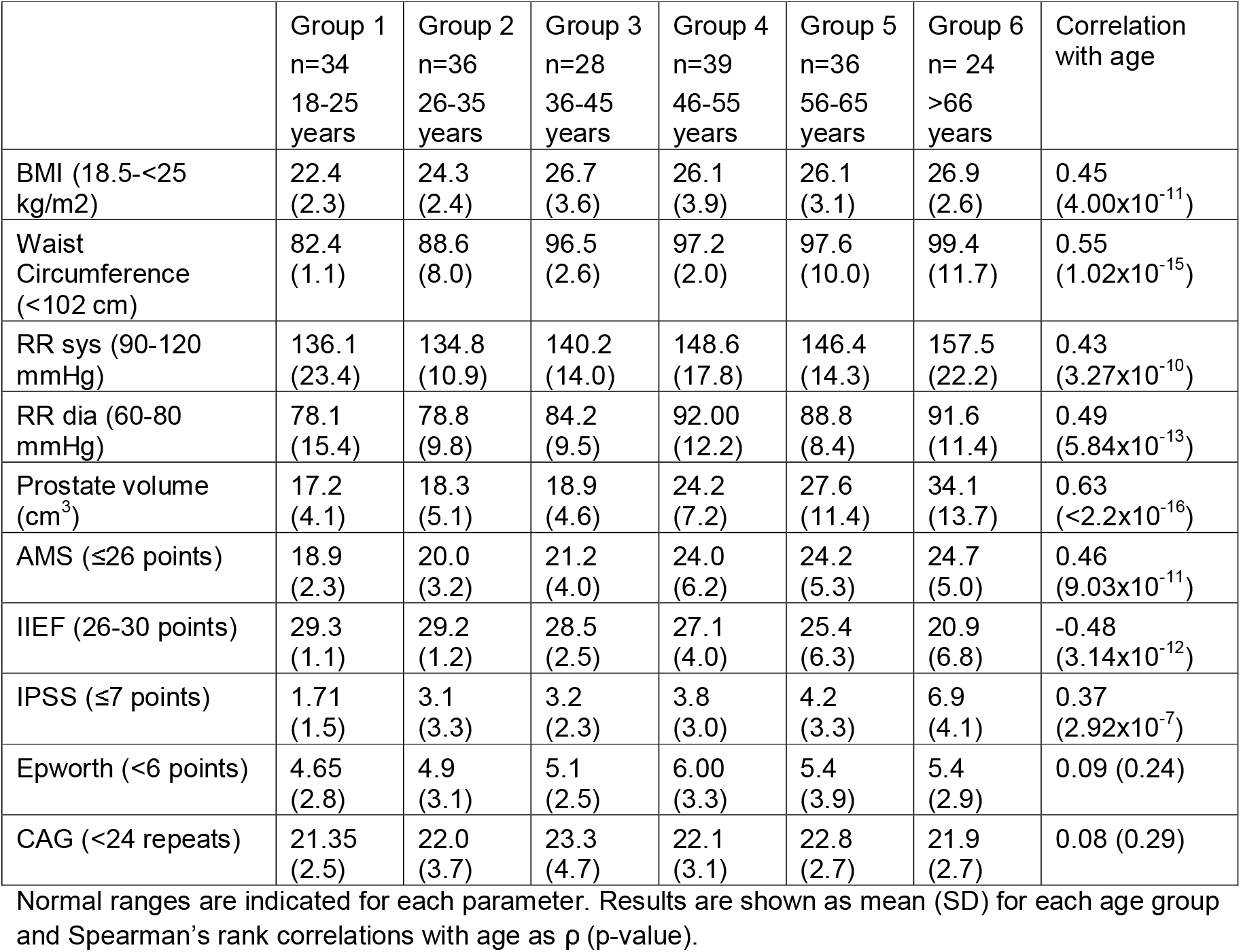
Anthropometric parameters and questionnaire results of the men in the cohort

|  | Group 1<br>n=34<br>18-25<br>years | Group 2<br>n=36<br>26-35<br>years | Group 3<br>n=28<br>36-45<br>years | Group 4<br>n=39<br>46-55<br>years | Group 5<br>n=36<br>56-65<br>years | Group 6<br>n= 24<br>>66<br>years | Correlation<br>with age |
| --- | --- | --- | --- | --- | --- | --- | --- |
| BMI (18.5-<25<br>kg/m<sup>2</sup>) | 22.4<br>(2.3) | 24.3<br>(2.4) | 26.7<br>(3.6) | 26.1<br>(3.9) | 26.1<br>(3.1) | 26.9<br>(2.6) | 0.45<br>(4.00x10<sup>-11</sup>) |
| Waist<br>Circumference<br>(<102 cm) | 82.4<br>(1.1) | 88.6<br>(8.0) | 96.5<br>(2.6) | 97.2<br>(2.0) | 97.6<br>(10.0) | 99.4<br>(11.7) | 0.55<br>(1.02x10<sup>-15</sup>) |
| RR sys (90-120<br>mmHg) | 136.1<br>(23.4) | 134.8<br>(10.9) | 140.2<br>(14.0) | 148.6<br>(17.8) | 146.4<br>(14.3) | 157.5<br>(22.2) | 0.43<br>(3.27x10<sup>-10</sup>) |
| RR dia (60-80<br>mmHg) | 78.1<br>(15.4) | 78.8<br>(9.8) | 84.2<br>(9.5) | 92.00<br>(12.2) | 88.8<br>(8.4) | 91.6<br>(11.4) | 0.49<br>(5.84x10<sup>-13</sup>) |
| Prostate volume<br>(cm<sup>3</sup>) | 17.2<br>(4.1) | 18.3<br>(5.1) | 18.9<br>(4.6) | 24.2<br>(7.2) | 27.6<br>(11.4) | 34.1<br>(13.7) | 0.63<br>(<2.2x10<sup>-16</sup>) |
| AMS (≤26 points) | 18.9<br>(2.3) | 20.0<br>(3.2) | 21.2<br>(4.0) | 24.0<br>(6.2) | 24.2<br>(5.3) | 24.7<br>(5.0) | 0.46<br>(9.03x10<sup>-11</sup>) |
| IIEF (26-30 points) | 29.3<br>(1.1) | 29.2<br>(1.2) | 28.5<br>(2.5) | 27.1<br>(4.0) | 25.4<br>(6.3) | 20.9<br>(6.8) | -0.48<br>(3.14x10<sup>-12</sup>) |
| IPSS (≤7 points) | 1.71<br>(1.5) | 3.1<br>(3.3) | 3.2<br>(2.3) | 3.8<br>(3.0) | 4.2<br>(3.3) | 6.9<br>(4.1) | 0.37<br>(2.92x10<sup>-7</sup>) |
| Epworth (<6 points) | 4.65<br>(2.8) | 4.9<br>(3.1) | 5.1<br>(2.5) | 6.00<br>(3.3) | 5.4<br>(3.9) | 5.4<br>(2.9) | 0.09 (0.24) |
| CAG (<24 repeats) | 21.35<br>(2.5) | 22.0<br>(3.7) | 23.3<br>(4.7) | 22.1<br>(3.1) | 22.8<br>(2.7) | 21.9<br>(2.7) | 0.08 (0.29) |

Normal ranges are indicated for each parameter. Results are shown as mean (SD) for each age group and Spearman's rank correlations with age as p (p-value).

### Molecular signatures of male germ cells in healthy ageing men

We investigated to what extent hallmarks of ageing previously established in somatic cells, such as telomere attrition, loss of genome stability, and epigenetic alterations, are affected in human sperm from ageing men.

We found an inverse association between age and blood rTL (r=-0.41, p=3.27×10-9), indicating the shortening of telomeres with age in somatic cells (Figure 1A). In contrast, there was a positive association of similar strength between age and sperm rTL (r=0.41, p=3.37×10-8), demonstrating that telomeres in the germline become longer with increasing age. Moreover, the rate at which telomeres shorten in blood DNA (0.0055 relative units per year) is slower than the rate that they lengthen in sperm (0.0077 relative units per year).

**Figure 1.**
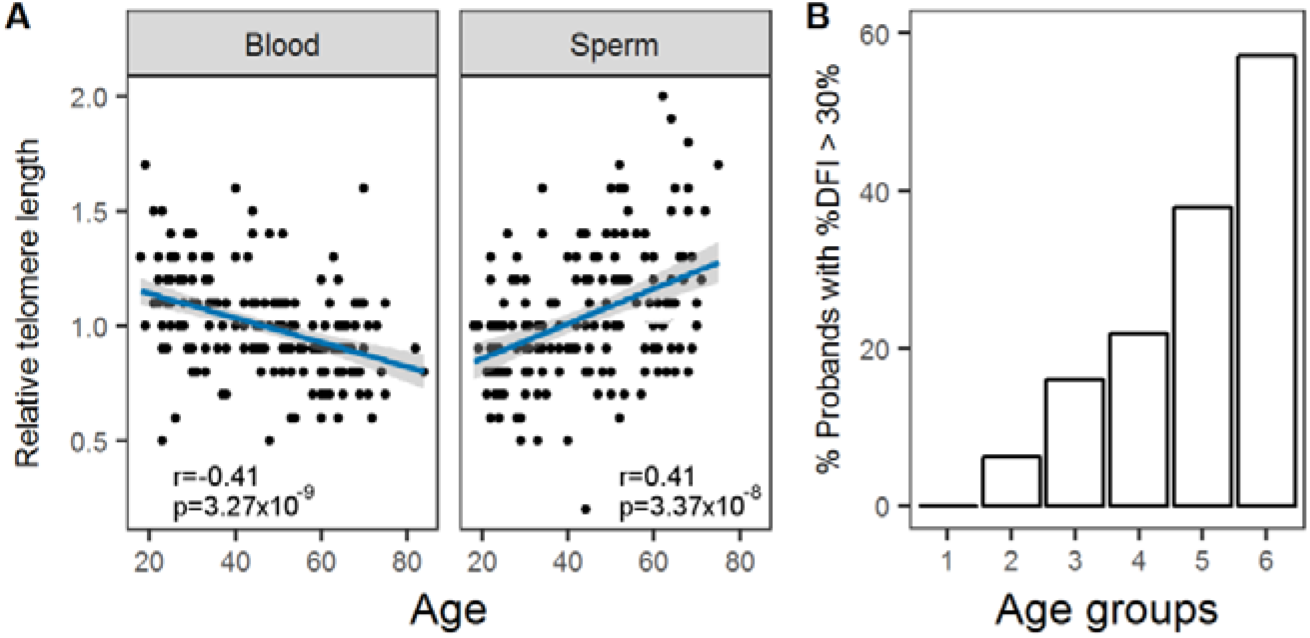
Relative Telomere Length (rTL) sperm DNA and sperm DNA fragmentation changes with age. A. rTL was measured in peripheral blood (left; n=194) and swim-up sperm (right; n=179) DNA. Linear trends are displayed in blue and 95% confidence interval in grey shading. The correlation coefficients (Pearson’s) and p values are indicated for each case. B. The percentage of men with a sperm DNA fragmentation index (DFI) higher than 30%, considered to negatively affect fertility, increased with age (n=169).

Genome instability was assessed in the paternal genome by measuring DNA fragmentation, in particular single- and double-strand DNA breaks (19). In the sperm of healthy men, we observed a highly significant increase in DFI with increasing age (ρ=0.57; p=1.04⨯10^−15^). Possible confounding parameters, such as abstinence time and sperm vitality, had little or no effect on this association (Table S1). While we found that the DFI continuously and steadily increased over the different age groups until the age of 55 years (Groups 1-4), a marked acceleration in genome instability was noted in the two oldest age groups (groups 5 and 6; Figure 1B), with almost 60% of men over the age of 66 showing DFI above 30%, which is considered the threshold for normal DFI (20).

Another hallmark of ageing seen in somatic cells is epigenetic drift, in which DNA methylation changes occur with age. As a proxy for assessing global DNA methylation in tissues or cells, we measured the DNA methylation of LINE-1 repetitive elements in bisulfite-converted blood DNA from our cohort, but did not find any age-related methylation changes (Figure S1).

The stability of LINE-1 methylation suggests that any age-dependent changes to the methylome might occur at unique regions in the genome. Using deep bisulfite sequencing (DBS), we assessed the sperm DNA methylation levels of regions previously found to be aberrantly methylated and associated with reduced fertility (21). These included two imprinted genes (*H19* and *MEST*) and four spermatogenesis-specific genes (*FGFR3, VASA, RHOXF1*, and *RHOXF2/2B*). In parallel, we also analysed blood DNA to exclude a general age-related tissue-independent effect. There were only minor differences detected between the younger (n=14, age range 18-25) and older (n=14; age range 61-71) age groups in DNA methylation levels of *H19* and *VASA* in blood DNA and in *FGFR3* in sperm DNA (Figure S2). It is therefore likely that age-dependent changes to the methylome are located at genomic loci not previously associated with male infertility.

In order to explore age-dependent methylation changes in sperm in an unbiased and genome-wide manner, we performed whole genome bisulfite sequencing (WGBS) on 6 samples each of sperm DNA from the two extreme age groups (Group 1 vs. Groups 5+6). For comparison, blood DNA from the same men were prepared and sequenced. There were no differences in mean DNA methylation between age groups or between tissues (Table S2; confirmation of purity of samples shown in Figure S3).

Differentially methylated regions (DMRs; with at least 4 CpGs and 30% mean methylation difference) between the extreme groups were identified using two different algorithms (BSmooth and Metilene (22, 23)). We identified a total of 236 DMRs (Supplementary File 1) in sperm and 36 in blood. Of the sperm DMRs, 121 increased and 115 decreased methylation with age. Notably, there was no overlap between the age-associated sperm DMRs and blood DMRs, consequently only the sperm DMRs were analysed further by bioinformatic and analytical tools. We did not find any chromosomal preference or hotspot for the observed sperm methylation changes (Figure S4).

Analysing the identified sperm DMRs, we noticed a significant overrepresentation of DNA transposons (p=9.59×10-4) and LINE elements (p=6.21×10-3), but no significant over- or underrepresentation of either LTRs or SINE (Figure S5).

A subset of 11 sperm DMRs was chosen for further in-depth analysis and used to confirm findings in samples from all of the age groups using DBS (Table S3). In addition to the 12 samples originally analysed by WGBS, 30-63 sperm samples, equally distributed throughout the six age groups, were randomly selected. By DBS analyses, we confirmed that seven out of eleven DMRs (BS21, BS74, BS134, ML15, BS71, and BS174) had a statistically significant correlation between DNA methylation levels and age. Two DMRs (BS46 and ML12) showed a similar trend as identified by WGBS, but the correlation with age was not statistically significant. Two other DMRs (BS88 and BS130) displayed a non-linear pattern of age-related methylation change (Figure 2).

**Figure 2.**
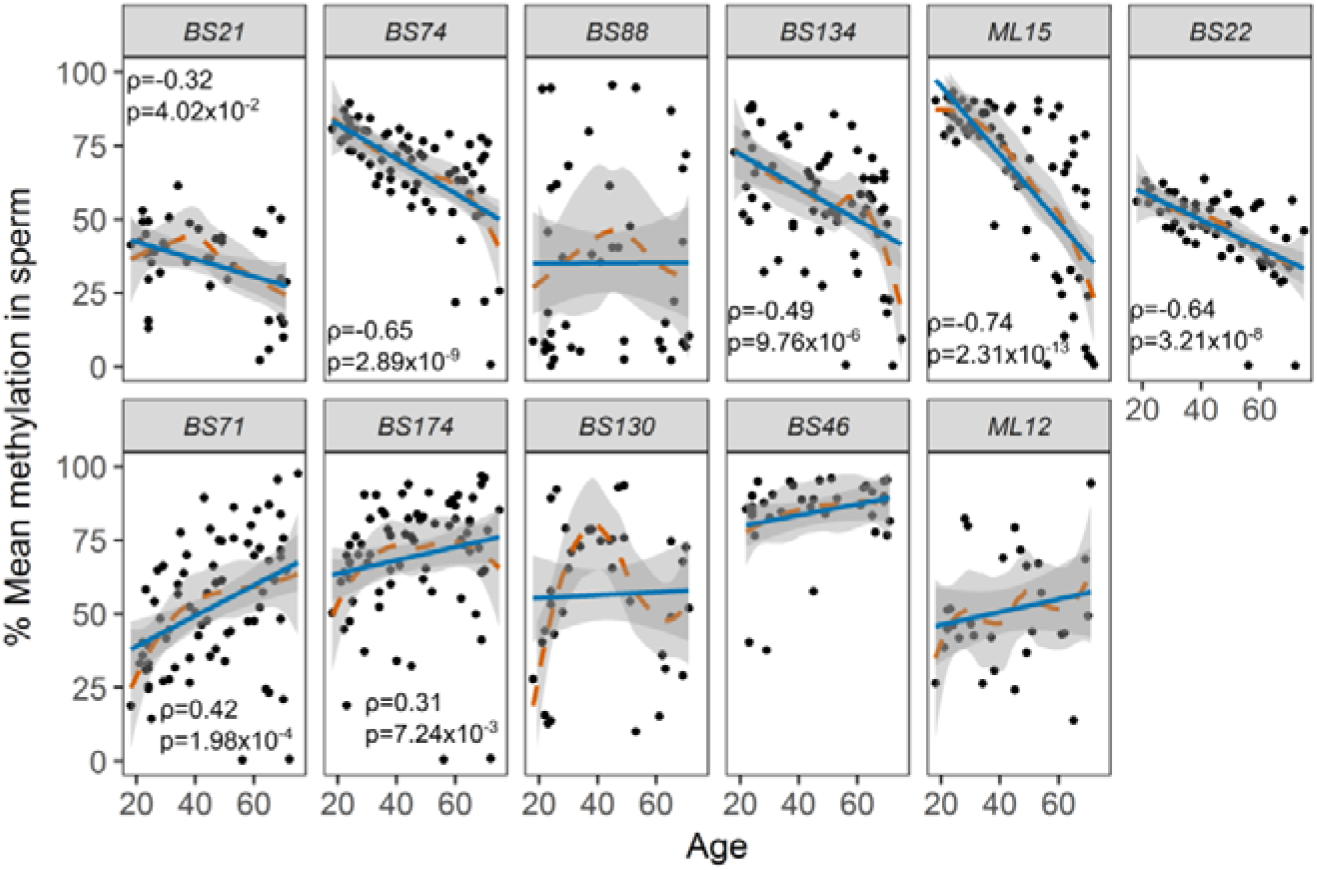
Evaluation of DMRs for age-dependent changes by DBS. Eleven age-associated differentially methylated regions (DMRs) identified by WGBS of sperm of young (G1) and old (G5-6) men were further analysed in all age groups (n≥7 for each group) by DBS. A statistically significant correlation (Spearman’s rank correlation) with age was detected for seven of these DMRs. Only statistically significant correlation results are displayed in each graph. The linear trend line is shown in blue and the LOWESS curve in dashed orange, with respective 95% confidence levels in grey shading.

The correlation between age and methylation of the different DMRs was adjusted for the possible confounder MTHFR C677T genotype, previously shown to influence DNA methylation. However, we could not observe an overall significant effect of this polymorphism on DNA methylation of the regions analysed.

We used six DMRs with p<0.001 (BS74, BS134, ML15, BS22, BS71 and BS174) and a regression analysis of donor-wise average methylation levels to derive an age predictor for sperm. By leave-one-out cross-validation (24), we found a strong correlation between predicted and chronological ages for the 42 initially analysed samples. An additional set of 33 samples were independently analysed, confirming this strong correlation (Figure 3; R script available in Supplementary File 2). Taking the two sample sets together, we found a strong correlation (r=0.78; p=3.83×10-16) between chronological and predicted age and a mean absolute error of 8.7 years.

**Figure 3.**
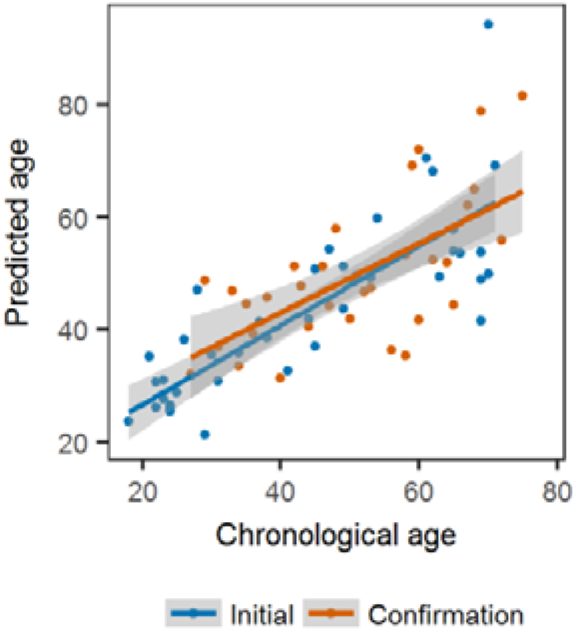
Sperm age predictor. The average methylation values of the six DMRs with a p<0.01 were used for regression analysis to develop an age-predictor. The predicted age, as determined by leave-one-out cross-validation, is plotted against the chronological age in blue for 12 probands from the WGBS discovery cohort and 30 additional probands, with the linear trend displayed in blue and 95% confidence interval in grey shading. An independent sample cohort (n=33) was analysed and the age predictor tool could be successfully applied to calculate the predicted age (results in orange). The calculated mean absolute error including all the samples was +/- 8.7 years, with a Pearson correlation coeficient of 0.78 (p=3.83×10^−16^).

In order to analyse the functional importance and potential biological impact of the 236 sperm DMRs, a list with the upstream and downstream neighbouring genes (distance from TSS ≤1000 kb) was obtained. Gene ontology (GO) analysis of the DMR-associated gene list showed a significant enrichment for Homeobox genes (16 genes; p_adj_=2.0×10^−2^) and other DNA binding protein gene families (58 genes; p_adj_=4.3×10^−2^). Gene set enrichment analysis showed enrichment in several gene sets (FDR<0.05), of which the ten most significant candidates (for the GO Biological Process Ontology) are highlighted in Figure 4.

**Figure 4.**
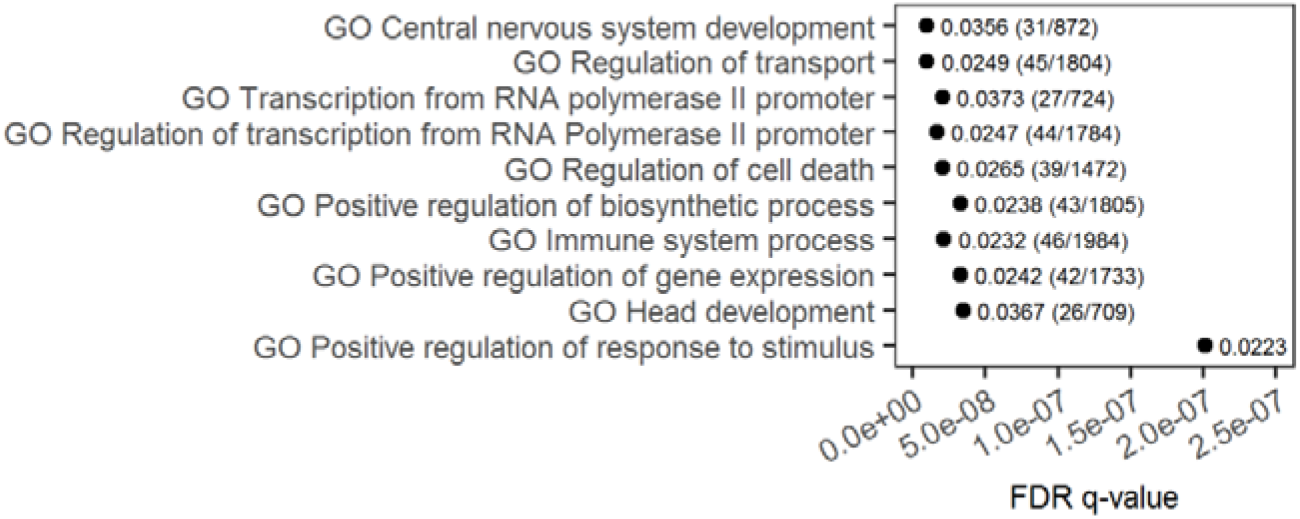
Biological impact of identified DMRs. Biological impact of identified DMRs. Overlap of the DMR-associated gene list with GO Biological Process, calculated using GSEA, is shown along with the false discovery rate (FDR) q-value. Only the 10 most significant pathways are displayed. For each term we display the ratio between the number of genes that overlap and the total number of genes in each gene enrichment set with the ratio components between brackets.

## Discussion

The FAMe study of healthy ageing men delineates for the first time an inherent ageing process that take place in germ cells during spermatogenesis by minimizing the effects of confounding factors. Considering the hallmarks of ageing already known for other tissues and cells, it is evident that male germ cells age, but the ageing process is different from that of somatic cells.

Good general health seems to be an essential requirement for good reproductive health, regardless of age, while severe male infertility appears to be associated with increased morbidities (25). Our andrological evaluation revealed that sperm production is not affected by age in a clinically relevant way. These findings contrast with several studies which showed a decline in endocrine and spermatogenic function with age (4, 6, 26). This is likely due to the strict health criteria applied during our volunteer recruitment, which were not used in most previous studies. Our results are in complete agreement with a previous study which also applied strong selection criteria (27). A possible limitation of this study is that we do not have longitudinal data, and therefore we cannot exclude an influence of the year of birth on the data.

We have found a negative correlation between age and blood rTL and a positive correlation of similar strength between age and sperm rTL, demonstrating an extension of telomeres in the germline with increasing age. This is presumably due to telomerase activity in the germline (28). The increased rate of telomere lengthening in sperm than telomere shortening in blood is in accordance with Aston et al. (29). There is evidence that very long sperm telomeres are detrimental to fertility (30). The age-dependent increase of telomeres in sperm could also result in increased telomere length in the progeny of elderly fathers; however, how this might affect their general health is currently unclear (31).

In our cohort, DFI increased with age, indicating increased genomic instability in the germline. Previous studies have shown that an increase in DFI above 30% is associated with male infertility (20). Consequently, the fertility of elderly men might be impaired by increased DNA instability, contributing to previously reported longer time to pregnancy and higer miscarriage rates irrespective of female partner age (32, 33). The number of men with DFI levels above 30% increased more dramatically in groups 5 and 6 (>56 years; Figure 1B), affecting close to 60% of the volunteers in the oldest group, pointing to an accelerated increase in DNA damage, a pattern of DNA instability hitherto unknown.

To our knowledge, this is the first study investigating age-related methylation changes in all CpG dinucleotides across the human sperm genome. We identified over 200 DMRs in sperm, none of which overlaped with DMRs identified in blood in this study or with any of the 353 CpGs from Horvath’s epigenetic clock (34). This strongly supports the notion that epigenetic ageing in the male germline differs from somatic cell ageing.

For a majority of the DMRs validated by DBS, a significant linear pattern of hyper- or hypomethylation with age was evident. The range of methylation changes observed was remarkable. For example, the mean methylation decreased from 87% to 46% (for ML15) and increased from 32% to 62% (for BS71) with age. Two candidate DMRs (BS88 and BS139) showed a non-linear pattern which might indivate a strong impact of other factors (e.g. genetic background). DNA methylation in sperm has been previously shown to be affected by genetic variants, such as *MTHFR* polymorphisms, however we could not find any influence of this variation on the DNA methylation levels of the analysed DMRs.

The inter-individual variation in DNA methylation was very low in most DMRs validated, which allowed us to devide an epigenetic age-predictor for sperm based on six of these regions. Recently, an epigenetic clock for sperm has been published by Jenkins et al. (16), which is more precise than our own. One reason for this difference may be the narrower age range in their cohort (20-60 years) and also that our predictor is based on only six regions. We think the two epigenetic clocks might complement each other, as they likely reflect different factors. Jenkins’ study cohort includes clinically heterogeneous sperm samples, including infertile patients, and therefore their clock may reflect not only the effect of age but also other factors such as age-related morbidities. In contrast, our data are based on a highly selected population of healthy men and therefore can be used for tracking the influence of health, fertility, lifestyle, and treatment on the ageing status of the germline.

Gene ontology analyses of genes in the vicinity of the identified sperm DMRs indicate an enrichment of biological processes involved in nervous system and head development (Figure 4). A similar finding was reported previously (15). Although it is tempting to speculate that there might be a link between this finding and the increased frequency of neuropsychiatric disorders in the offspring of older men, there is currently no experimental support for such a hypothesis.

A crucial question emerges from our study: what is the origin of the epigenetic changes in sperm? It is unlikely that DNA methylation is altred during spermiogenesis, where DNA is tightly packed with protamines and therefore inaccessible for transcription factor binding and modification by enzymes. The spermatogenial stem cells migh be a source of the observed changes. At 60 years of age, it is estimated that a spermatogonia has undergone approximately 920 mitotic cell divisions (35), making them prone to replication errors and resulting in increased mutation rates. It is therefore likely that male germ cells are also prone to increased DNA methylation maintenance errors as they age. Moreover, to maintain sperm production levels, testes of older men require require activation and recruitment of additional, otherwise quiescent, spermatogonia (36). This changed spermatogonial activity pattern could cause the observed methylation, and even DNA instability, changes. In addition, similarly to what happens with mutations (35), certain DNA methylation changes might confer a selective advantage to some spermatogonia, which would contribute more to the spermatogenic output with increasing age. This would indicate that two further hallmarks of ageing, spermatogonial stem cell replication stress and clonal drift, could contribute significantly to the ageing of the male germline.

In summary, healthy ageing is strongly associated with good reproductive health, with normal semen parameters being maintained over a six decades-long period. However, this starkly contrasts with intrinsic ageing processes occurring in the male germline, such as telomere lengthening, increasing genome instability, and DNA methylation changes at specific sites. These molecular changes might contribute to the reduced fertility and fecundity in older men.

It has been previously advised that genetic counselling should be provided for couples undergoing ART when the male partner is over 49 years of age (37, 38). Our data support this recommendation, but further work will be necessary in the future to understand whether the molecular changes to sperm DNA have consequences to progeny health, to set precise age thresholds, and ultimately whether fertility preservation options (e.g. sperm cryopreservation) should be offered to men while they are still young.

## Materials and Methods

### Ethics

All participants gave informed written consent for performing the examinations, evaluation of clinical data and genetic analysis of the DNA samples according to protocols approved by the Ethics Committee of the Medical Faculty in Münster (2013-255-f-S).

### Participants

Participants were recruited through advertisements in local newspapers, internet, and hospitals. The recruitment and clinical evaluation/selection of the participants took place between October of 2014 and April of 2016. Prospective participants (above the age of 18) were first asked to fill an online questionnaire. Exclusion criteria in the questionnaire included smoking within the previous year, illegal drug consumption, regular use of medication (except for the treatment of mild hypertension, hypothyroidism, and dyslipoproteinemia), hospitalization within the previous month, current or former cancer/cancer treatment, severe chronic renal failure, chronic viral infection, urogenital malformations and surgeries, prior diagnosis or treatment for fertility impairment, chromosomal alterations, and participation in clinical trials within the previous year. 265 selected volunteers were invited to the outpatient clinic of the CeRA for clinical evaluation and sample collection. 65 volunteers dropped out for unwillingness or inability to make it to the appointment. 200 men underwent a thorough anthropometric, endocrine, and andrological evaluation, which was accompanied by several questionnaires addressing their wellbeing and sexual health. Two participants were further excluded due to the discovery of assymptomatic tumours during the clinical examination. 197 participants were ultimately included in the study.

Physical examination included measurement of body weight, height, blood pressure, pulse, and electrocardiogram. Blood samples were collected under fasting conditions and used for DNA isolation, hormone measurements, haematological analysis including metabolic parameters as blood lipids, HbA1c and blood glucose. Ultrasound of the thyroid, carotid arteries, kidneys, bladder, testicles, epididymis, prostate, and seminal vesicles (transrectal) in addition to palpation of breast, external genitalia, and digital rectal examination were performed. The volunteers were also asked to provide at least 2 semen samples in two separate visits, with an abstinence time of 2 -7 days. The semen samples were analysed in our andrological lab in accordance with the WHO criteria (18). A microbiological analysis of semen was performed and swim-up sperm was prepared (18) and stored for further analyses. All collected data were stored in our in-house clinical database, Androbase (39).

Grouping was done in 10 year intervals, except for groups 1 (18-24 years) and 6 (> 65 years; range 65 - 82 years), the latter due to the difficulties in recruiting sufficient numbers of healthy men above 75 years of age (Table 1).

### Questionnaires

General wellbeing including personal medical history, life time smoking habits, socio-economic factors, and family medical history were analysed using four different questionnaires. Clinical symptoms of age-related hypogonadism were evaluated using the Aging Males Symptoms (AMS) rating scale (40). Erectile function was assessed with the international index of erectile function (IIEF) (41, 42). The evaluation of clinical symptoms caused by an enlargement of the prostate was measured by the International Prostate Symptom Score questionnaire (43). Quality of daily sleep was assessed by the Epworth sleepiness scale (44).

### Hormone measurements

Hormonal parameters (LH, FSH, T, SHBG, prolactin, estradiol, PSA) were measured using a chemiluminescent microparticle immunoassay (Architect i1000; Abbott Diagnostics, Wiesbaden, Germany). DHT was measured using an in-house radio-immunoassay. All blood samples were obtained in a fasting state after 15 minutes of rest. Methods are validated quarterly against LC-MS/MS.

### DNA Fragmentation Index (DFI) determination

Measurement of DNA fragmentation was performed as described previously (45). For the flow cytometer setup and calibration, a reference sample was used from a normal donor. Data were analysed using the FCS 3.0 software package (DeNovo software, Los Angeles, CA, USA). A DNA fragmentation index (DFI) for every detected sperm was calculated according to the formula: red/(red + green) fluorescence. DFI values were plotted in a histogram and sperm with denatured DNA (high DFI values) were those located to the right of the main, normal population.

### DNA isolation and bisulfite conversion

Blood DNA was isolated using EDTA-blood using the FlexiGene DNA kit (Qiagen) according to the instructions supplied by the manufacturer and as previously described (46). Sperm DNA was isolated using MasterPure DNA purification kit (Epicentre Biotechnologies, Madison, WI, USA) and a modified protocol (46). DNA concentration was measured by spectrophotometry (NanoDrop ND-1000, Peqlab, Erlangen, Germany). DNA samples were stored at -20°C until further use.

For LINE-1 DNA methylation determination, DNA was bisulfite converted using the EpiTect kit (Qiagen). Briefly, 200 ng blood DNA were bisulfite converted using Qiagen EpiTect according to the manufacturer’s instructions. The converted DNA was eluted in 20 μL Buffer EB.

For Deep Bisulfite Sequencing (DBS), 200 ng blood and sperm DNA were bisulfite converted using the EZ DNA Methylation Gold kit (Zymo Research, Freiburg, Germany) according to the provided instructions, and the DNA was eluted in 10 μL.

### Relative Telomere Length Determination

Mean relative telomere length (rTL) was measured on blood and swim-up sperm DNA by quantitative PCR (qPCR) using a previously described method (47), modified from (48).

Briefly, the cycle threshold (Ct) value for telomeric repeats (T) and a single-copy gene [S; haemoglobin subunit gamma (*HBG*)] were determined for each sample and the rTL was calculated as the ratio between T and S. A standard curve was generated using reference DNA ranging from 0.37 to 90 ng from 10 randomly selected subjects. Measurements for subjects and reference samples were carried out in triplicates. Amplification mixture contained 10 µl QuantiTect SYBR^®^ green PCR Master Mix (2x; Qiagen), 2 µl blood DNA (12,5 ng/μl), 1 µl primer either for telomeric repeats (forward 2 µM, reverse 18 µM; Eurofins MWG) or *HBG* (forward 6 µM, reverse 14 µM; Eurofins MWG; Table S4, 2 µl DTT (25 mM), and nuclease free water to a total volume of 20 µl.

### LINE-1 methylation determination

The DNA methylation of LINE-1 repetitive elements was quantified using PyroMark Q24 CpG LINE-1 kit (Qiagen) according to the procedure advised by the manufacturer.

### Library preparation and sequencing

Sperm and blood DNA, respectively, from six “young” men (18-24 years) and six “old” men (61-71 years) were pooled at equimolar ratios. We used pooled DNA to minimize the effect of inter-individual DNA methylation differences (49–51). For each of the 4 pools, two shotgun libraries were generated. Standard pre-bisulfite libraries were prepared from 2 μg of DNA according to the original Illumina protocol essentially as described by Rademacher et al. (52). Tagmentation libraries were prepared from 20 ng of DNA using the Nextera transposase (Illumina, San Diego, CA, USA) and a modified protocol (personal communication from G. Gasparoni and J. Walter, University of Saarland). Bisulfite conversion was performed with the EZ DNA Methylation-Gold Kit (Zymo Research, Irvine, CA, USA), followed by 10 and 12 cycles of PCR, respectively. For each sample, we sequenced two lanes of the Illumina library and one lane of the tagmentation library on an Illumina HiSeq 2500, again as described previously (52).

Whole genome bisulfite sequencing (WGBS) data was deposited at the European Nucleotide Archive under the accession number PRJEB28044.

### Read mapping and methylation calling

Raw reads were aligned with bwa-meth (v0.2.0) (53) to the human reference genome (hs37d5) with default parameters. Aligned reads were sorted and indexed, duplicates were detected, and alignment metrics were extracted by sambamba (v.0.6.6) (54). Methylation levels at CpG dinucleotides were called by an in-house script, which considers only CpGs located on the reference genome.

### DMR detection

Differentially methylated regions (DMRs) were detected by Metilene (v.0.2-7) (23) and by an in-house script based on Bsmooth (22) as they detect preferentially shorter and longer DMRs, respectively. We removed the smoothing and detect differentially methylated CpGs (DMCs) by t-test as previously described (22). Adjacent differentially methylated cytosines (DMCs) are merged into a DMR if they are separated by not more than two non-DMCs. DMRs containing four DMCs and a minimum of 0.3 (i.e. 30%) average group methylation difference between these DMCs were selected.

### Deep Bisulfite Sequencing (DBS)

Primer sets (Table S4) were designed for a selection of the detected DMRs using MethPrimer (55) (http://www.urogene.org/cgi-bin/methprimer/methprimer.cgi) and MethPrimer 2.0 (http://www.urogene.org/cgi-bin/methprimer2/MethPrimer.cgi). Only DMRs not overlaping with repetitive sequences were selected, in order to generate highly specific primer sequences. DMR-specific libraries were constructed by two rounds of PCR as previously described (46, 56). Sample preparation and sequencing were performed as previously described (56) using the Roche/454 GS Junior system (Roche Diagnostics, Manheim, Germany) and yield of reads was increased by applying a special filter setting (57). A power analysis was performed to find the minimum number of samples to be analysed (p<0.05 and β<0.2, large effect size).

Methylation analysis was performed using Amplikyzer (58), and average methylation for each amplicon and each CpG site, as well as methylation plots, were retrieved for each sample.

### Gene Ontology Analysis

Annotation of the DMRs was performed by searching the nearest genes within 100 kb. The resulting gene list was submitted to DAVID (59) was used for gene ontology, using the whole human genome as backgroun. GSEA (60) was used for gene set enrichment analysis using the Molecular Signatures Database (MsigDB).

### Overlap of DMRs with repeats

Repeat tracks produced by RepeatMasker (61) were retrieved by the UCSC Table Browser (62). Separated BED files for the four different types of repeats were generated and intersected with CpG positions. This results in four BED files, each containing CpGs located in the respective repeat regions. CpGs of DMRs located in repeat regions were then identified by intersecting the DMRs in BED format with each of the four files. All operations on BED files are performed by BEDTools (v2.25.0) (63).

We estimated the p-value for over- and underrepresented repeat overlaps by simulating 1 million datasets, consisting of 236 regions each and equal size distribution compared to our DMRs, while each region contains at least 4 CpGs. For each of our 236 DMRs and each region of the simulated datasets, the fraction of repeat-overlapping CpGs was detected. We then compared the mean fraction f for our DMRs and each of the 1 million random sets F’. The empirical p-value for overrepresentation is the fraction of random sets f’ ∈ F’ with f > f’ (symmetrically, for underrepresentation). This was performed independently for each type of repeat.

### Epigenetic age-predictor

Let *X* be the set of Donors, *D* our six selected DMRs, *m*_*x*_*(d)* be the average methylation level (if available) for DMR *d* ∈ *D* and donor *x* ∈ *X* and *a(x)* its age.

We use the set *T*_*i,j*_ *= {(m*_*x*_*(d*_*i*_*), m*_*x*_*(d*_*j*_), *a(x))* for *x* ∈ *X*} to separately train a ridge regression (64) predictor *p*_*i,j*_ for each pair *(d*_*i*,_ *d*_*j*_) ∈ *D^2^* with *0 <= i < j <* |*D*|, while elements with at least one missing methylation value for *x* are removed from *T*_*i,j*_.

Each *p*_*i,j*_ is able to predict an age *a’*_*i,j*_*(x) = p*_*i,j*_*(m*_*x*_*(d*_*i*_*), m*_*x*_*(d*_*j*_)) given average methylation of *x* for *d*_*i*_ and *d*_*j*._ Let then a’(x) be the average of all single predictions for *x*.

In a second step we use all pairs *T’*_*i,j*_ *= {(a’(x), a(x)) for x* ∈ *X}* to train an additional simple linear regression model *q*. The final predicted age for *x* is is *q(a’(x))*.

### Statistics

For each analysed variable, normality of the distribution and homogeneity of variance were tested before determining the most adequate statistical test to use. Two-sided t-tests were used to compare betweens between two groups. Pearson’s test was used to test correlations between age and normally distributed variables. Spearman’s rank correlation was used when the variables were not normally distributed or presented outliers. Adjustment for confounders was done by partial correlation analysis. All statistical analysis and graph plotting were performed using R 3.3.1 and suitable R packages.

## Data Availability

WGBS data was deposited at the European Nucleotide Archive under the accession number PRJEB28044.

## Acknowledgments

We would like to thank all men who contributed as probands in this study. We thank Sabine Forsthoff, Claudia Haak, Daniela Hanke, Jolanta Körber, Raphaele Kürten, Elisabeth Lahrmann, Susanne Orlowski, Sonja Barkhaus, Elena Plester, Reinhild Sandhowe, Sabine Strüwing, and Nicole Terwort for technical assistance, Ludger Klein-Hitpass for Illumina sequencing and Elsa Leitão and Michael Zeschnigk for helpful discussions.

## Author contributions

S.L. did the experimental study design, sample and data analyses, evaluated clinical parameters, and wrote the manuscript. J.F.C. did the clinical study design, proband recruitment, and data analyses. B.H. did the epigenetic study design and WGBS analyses, data evaluation, and contributed to the manuscript. F.T. contributed to the experimental and clinical study design and the manuscript. K.C. worked on the study design, online questionaire, the ethical vote, and proband recruitment. M.Z. worked on the study design, proband recruitment and examination. E.P. did experimental work, was involved in data interpretation, and worked on the manuscript. S.R. performed the bioinformatics analyses of the WGBS data. C.S. set up the epigenetic clock and did WGBS analyses. S.B. performed DMR analyses. K.R. did the DFI analysis. C.K. worked on the study design, proband recruitment and examination. St.S. worked on the study design, analysed the data and worked on the manuscript. S.K. was responsible for the clinical study design, the ethical permission, and proband examination. J.G. developed the concept of the study, performed clinical and experimental data analyses, and wrote the manuscript.

## Data availability

WGBS data was deposited at the European Nucleotide Archive under the accession number PRJEB28044.

## References

1. Mills M, Rindfuss RR, McDonald P, te Velde E. Why do people postpone parenthood? Reasons and social policy incentives. Hum Reprod Update 2011; 17(6):848–60.

2. Prioux F. Late fertility in Europe: some comparative and historical data. Rev Epidemiol Sante Publique 2005; 53 Spec No 2:2S3-11.

3. Khandwala YS, Zhang CA, Lu Y, Eisenberg ML. The age of fathers in the USA is rising: an analysis of 168 867 480 births from 1972 to 2015. Hum Reprod 2017; 32(10):2110–6.

4. Eskenazi B, Wyrobek AJ, Sloter E, Kidd SA, Moore L, Young S et al. The association of age and semen quality in healthy men. Hum Reprod 2003; 18(2):447–54.

5. Kelsey TW, Li LQ, Mitchell RT, Whelan A, Anderson RA, Wallace WHB. A validated age-related normative model for male total testosterone shows increasing variance but no decline after age 40 years. PLoS ONE 2014; 9(10):e109346.

6. Sartorius GA, Nieschlag E. Paternal age and reproduction. Hum Reprod Update 2010; 16(1):65–79.

7. Zitzmann M. Effects of age on male fertility. Best Practice & Research Clinical Endocrinology & Metabolism 2013; 27(4):617–28. Available from: URL: http://www.sciencedirect.com/science/article/pii/S1521690X13001127.

8. La Rochebrochard E de, Thonneau P. Paternal age and maternal age are risk factors for miscarriage; results of a multicentre European study. Hum Reprod 2002; 17(6):1649–56.

9. Koh S-A, Sanders K, Deakin R, Burton P. Male age negatively influences clinical pregnancy rate in women younger than 40 years undergoing donor insemination cycles. Reprod Biomed Online 2013; 27(2):125–30.

10. Mutsaerts MAQ, Groen H, Huiting HG, Kuchenbecker WKH, Sauer PJJ, Land JA et al. The influence of maternal and paternal factors on time to pregnancy--a Dutch population-based birth-cohort study: the GECKO Drenthe study. Hum Reprod 2012; 27(2):583–93.

11. Khandwala YS, Baker VL, Shaw GM, Stevenson DK, Lu Y, Eisenberg ML. Association of paternal age with perinatal outcomes between 2007 and 2016 in the United States: population based cohort study. BMJ 2018; 363:k4372.

12. Kong A, Frigge ML, Masson G, Besenbacher S, Sulem P, Magnusson G et al. Rate of de novo mutations and the importance of father’s age to disease risk. Nature 2012; 488(7412):471–5.

13. Maretty L, Jensen JM, Petersen B, Sibbesen JA, Liu S, Villesen P et al. Sequencing and de novo assembly of 150 genomes from Denmark as a population reference. Nature 2017; 548(7665):87–91.

14. Kovac JR, Addai J, Smith RP, Coward RM, Lamb DJ, Lipshultz LI. The effects of advanced paternal age on fertility. Asian J Androl 2013; 15(6):723–8.

15. Jenkins TG, Aston KI, Pflueger C, Cairns BR, Carrell DT. Age-associated sperm DNA methylation alterations: possible implications in offspring disease susceptibility. PLoS Genet 2014; 10(7):e1004458.

16. Jenkins TG, Aston KI, Cairns B, Smith A, Carrell DT. Paternal germ line aging: DNA methylation age prediction from human sperm. BMC Genomics 2018; 19.

17. Milekic MH, Xin Y, O’Donnell A, Kumar KK, Bradley-Moore M, Malaspina D et al. Age-related sperm DNA methylation changes are transmitted to offspring and associated with abnormal behavior and dysregulated gene expression. Mol Psychiatry 2015; 20(8):995–1001.

18. World Health Organization. WHO laboratory manual for the examination and processing of human semen. 5th ed. Geneva: World Health Organization; 2010.

19. Evenson DP. The Sperm Chromatin Structure Assay (SCSA(®)) and other sperm DNA fragmentation tests for evaluation of sperm nuclear DNA integrity as related to fertility. Anim Reprod Sci 2016; 169:56–75.

20. Evenson DP, Jost LK, Marshall D, Zinaman MJ, Clegg E, Purvis K et al. Utility of the sperm chromatin structure assay as a diagnostic and prognostic tool in the human fertility clinic. Hum Reprod 1999; 14(4):1039–49.

21. Laurentino S, Borgmann J, Gromoll J. On the origin of sperm epigenetic heterogeneity. Reproduction 2016; 151(5):R71–8.

22. Hansen KD, Langmead B, Irizarry RA. BSmooth: from whole genome bisulfite sequencing reads to differentially methylated regions. Genome Biol 2012; 13(10):R83.

23. Jühling F, Kretzmer H, Bernhart SH, Otto C, Stadler PF, Hoffmann S. metilene: fast and sensitive calling of differentially methylated regions from bisulfite sequencing data. Genome Res 2016; 26(2):256–62.

24. Webb GI, Sammut C, Perlich C, Horváth T, Wrobel S, Korb KB et al. Leave-One-Out Cross-Validation. In: Sammut C, Webb GI, editors. Encyclopedia of machine learning. New York, N.Y., London: Springer; 2011. p. 600–1.

25. Latif T, Kold Jensen T, Mehlsen J, Holmboe SA, Brinth L, Pors K et al. Semen Quality as a Predictor of Subsequent Morbidity: A Danish Cohort Study of 4,712 Men With Long-Term Follow-up. Am J Epidemiol 2017; 186(8):910–7.

26. Lee DM, O’Neill TW, Pye SR, Silman AJ, Finn JD, Pendleton N et al. The European Male Ageing Study (EMAS): design, methods and recruitment. Int J Androl 2009; 32(1):11–24.

27. Nieschlag E, Lammers U, Freischem CW, Langer K, Wickings EJ. Reproductive functions in young fathers and grandfathers. J Clin Endocrinol Metab 1982; 55(4):676–81.

28. Ozturk S. Telomerase Activity and Telomere Length in Male Germ Cells. Biol Reprod 2015; 92(2). Available from: URL: https://academic.oup.com/biolreprod/article-pdf/92/2/53,1-11/10556753/biolreprod0053.pdf.

29. Aston KI, Hunt SC, Susser E, Kimura M, Factor-Litvak P, Carrell D et al. Divergence of sperm and leukocyte age-dependent telomere dynamics: implications for male-driven evolution of telomere length in humans. Mol Hum Reprod 2012; 18(11):517–22.

30. Cariati F, Jaroudi S, Alfarawati S, Raberi A, Alviggi C, Pivonello R et al. Investigation of sperm telomere length as a potential marker of paternal genome integrity and semen quality. Reprod Biomed Online 2016; 33(3):404–11.

31. Eisenberg DTA, Kuzawa CW. The paternal age at conception effect on offspring telomere length: mechanistic, comparative and adaptive perspectives. Philos Trans R Soc Lond B, Biol Sci 2018; 373(1741).

32. Kennedy C, Ahlering P, Rodriguez H, Levy S, Sutovsky P. Sperm chromatin structure correlates with spontaneous abortion and multiple pregnancy rates in assisted reproduction. Reprod Biomed Online 2011; 22(3):272–6.

33. Spanò M, Bonde JP, Hjøllund HI, Kolstad HA, Cordelli E, Leter G. Sperm chromatin damage impairs human fertility. The Danish First Pregnancy Planner Study Team. Fertil Steril 2000; 73(1):43–50.

34. Horvath S, Raj K. DNA methylation-based biomarkers and the epigenetic clock theory of ageing. Nature Reviews Genetics 2018; 19(6):371–84.

35. Goriely A. Decoding germline de novo point mutations. Nat Genet 2016; 48(8):823–4.

36. Pohl E, Höffken V, Schlatt S, Kliesch S, Gromoll J, Wistuba J. Ageing in men with normal spermatogenesis alters spermatogonial dynamics and nuclear morphology in Sertoli cells. Andrology 2019.

37. Jennings MO, Owen RC, Keefe D, Kim ED. Management and counseling of the male with advanced paternal age. Fertil Steril 2017; 107(2):324–8.

38. Toriello HV, Meck JM. Statement on guidance for genetic counseling in advanced paternal age. Genet Med 2008; 10(6):457–60.

39. Tüttelmann F, Luetjens CM, Nieschlag E. Optimising workflow in andrology: a new electronic patient record and database. Asian J Androl 2006; 8(2):235–41.

40. Heinemann LAJ, Saad F, Zimmermann T, Novak A, Myon E, Badia X et al. The Aging Males’ Symptoms (AMS) scale: update and compilation of international versions. Health Qual Life Outcomes 2003; 1:15.

41. Rosen RC, Riley A, Wagner G, Osterloh IH, Kirkpatrick J, Mishra A. The international index of erectile function (IIEF): a multidimensional scale for assessment of erectile dysfunction. Urology 1997; 49(6):822–30.

42. Rhoden EL, Telöken C, Sogari PR, Vargas Souto CA. The use of the simplified International Index of Erectile Function (IIEF-5) as a diagnostic tool to study the prevalence of erectile dysfunction. Int J Impot Res 2002; 14(4):245–50.

43. Barry MJ, Fowler FJ, O’Leary MP, Bruskewitz RC, Holtgrewe HL, Mebust WK et al. The American Urological Association symptom index for benign prostatic hyperplasia. The Measurement Committee of the American Urological Association. J Urol 1992; 148(5):1549-57; discussion 1564.

44. Johns MW. A new method for measuring daytime sleepiness: the Epworth sleepiness scale. Sleep 1991; 14(6):540–5.

45. Evenson D, Jost L. Sperm chromatin structure assay for fertility assessment. Curr Protoc Cytom 2001; Chapter 7:Unit 7.13.

46. Laurentino S, Beygo J, Nordhoff V, Kliesch S, Wistuba J, Borgmann J et al. Epigenetic germline mosaicism in infertile men. Hum Mol Genet 2015; 24(5):1295–304.

47. Meyer A, Salewsky B, Buchmann N, Steinhagen-Thiessen E, Demuth I. Relative Leukocyte Telomere Length, Hematological Parameters and Anemia - Data from the Berlin Aging Study II (BASE-II). Gerontology 2016; 62(3):330–6.

48. Cawthon RM. Telomere measurement by quantitative PCR. Nucleic Acids Res 2002; 30(10):e47.

49. Cheung WA, Shao X, Morin A, Siroux V, Kwan T, Ge B et al. Functional variation in allelic methylomes underscores a strong genetic contribution and reveals novel epigenetic alterations in the human epigenome. Genome Biol 2017; 18(1):50.

50. Do C, Lang CF, Lin J, Darbary H, Krupska I, Gaba A et al. Mechanisms and Disease Associations of Haplotype-Dependent Allele-Specific DNA Methylation. Am J Hum Genet 2016; 98(5):934–55.

51. Schröder C, Leitão E, Wallner S, Schmitz G, Klein-Hitpass L, Sinha A et al. Regions of common inter-individual DNA methylation differences in human monocytes: genetic basis and potential function. Epigenetics Chromatin 2017; 10(1):37.

52. Rademacher K, Schröder C, Kanber D, Klein-Hitpass L, Wallner S, Zeschnigk M et al. Evolutionary origin and methylation status of human intronic CpG islands that are not present in mouse. Genome Biol Evol 2014; 6(7):1579–88.

53. Pedersen BS, Eyring K, De S, Yang IV, Schwartz DA. Fast and accurate alignment of long bisulfite-seq reads; 2014 Jan 6. Available from: URL: http://arxiv.org/pdf/1401.1129v2.

54. Tarasov A, Vilella AJ, Cuppen E, Nijman IJ, Prins P. Sambamba: fast processing of NGS alignment formats. Bioinformatics 2015; 31(12):2032–4.

55. Li L-C, Dahiya R. MethPrimer: designing primers for methylation PCRs. Bioinformatics 2002; 18(11):1427–31.

56. Beygo J, Ammerpohl O, Gritzan D, Heitmann M, Rademacher K, Richter J et al. Deep bisulfite sequencing of aberrantly methylated loci in a patient with multiple methylation defects. PLoS ONE 2013; 8(10):e76953.

57. Beygo J, Citro V, Sparago A, Crescenzo A de, Cerrato F, Heitmann M et al. The molecular function and clinical phenotype of partial deletions of the IGF2/H19 imprinting control region depends on the spatial arrangement of the remaining CTCF-binding sites. Hum Mol Genet 2013; 22(3):544–57.

58. Rahmann S, Beygo J, Kanber D, Martin M, Horsthemke B, Buiting K. Amplikyzer: Automated methylation analysis of amplicons from bisulfite flowgram sequencing. PeerJ Preprints 2013.

59. Huang DW, Sherman BT, Lempicki RA. Systematic and integrative analysis of large gene lists using DAVID bioinformatics resources. Nat Protoc 2009; 4(1):44–57.

60. Subramanian A, Tamayo P, Mootha VK, Mukherjee S, Ebert BL, Gillette MA et al. Gene set enrichment analysis: a knowledge-based approach for interpreting genome-wide expression profiles. Proc Natl Acad Sci U S A 2005; 102(43):15545–50.

61. Tarailo-Graovac M, Chen N. Using RepeatMasker to identify repetitive elements in genomic sequences. Curr Protoc Bioinformatics 2009; Chapter 4:Unit 4.10.

62. Karolchik D, Hinrichs AS, Furey TS, Roskin KM, Sugnet CW, Haussler D et al. The UCSC Table Browser data retrieval tool. Nucleic Acids Res 2004; 32(Database issue):D493–6.

63. Quinlan AR, Hall IM. BEDTools: a flexible suite of utilities for comparing genomic features. Bioinformatics 2010; 26(6):841–2.

64. Hoerl AE, Kennard RW. Ridge Regression: Applications to Nonorthogonal Problems. Technometrics 1970; 12(1):69.

